# Duration of Workplace Noise Exposure and Blood Pressure among Rural Adult Weavers

**DOI:** 10.1101/2025.05.23.25328248

**Authors:** Golam Dastageer Prince, Rajan Talukder, Obaidullah Ibn Raquib, Sharmim Akter, Dilkhush Jahan, Md. Rahul Parvez, Salah Uddin Ahmed, Sifatul Farid

**Author notes:** Corresponding author: Golam Dastageer Prince, MBBS, MPH.

## Abstract

This study aimed to determine the effect of duration of noise exposure on blood pressure among rural adult powerloom weavers. A cross-sectional study was done among 289 adult workers of selected weaving factories of Araihazar, Narayanganj, Bangladesh from January to December 2023. A semi-structured questionnaire, containing socio-demographic data, behavioral factors, family history, and disease history was developed as a data collection tool and data was collected by face-to-face interview. Blood pressure, height, weight, and noise intensity were measured accordingly.Among participants, 94.1% were male, 84.4% were married, and more than one-third were without formal education; their mean age was 34.41±10.17. The respondents’ mean duration of workplace exposure was 15.97 years and the measured intensity of noise inside the factories was 105.45 dB (96-111 dB). The mean systolic blood pressure (SBP) was 124.52 mmHg and the mean diastolic blood pressure (DBP) was 82.93 mmHg. The prevalence of hypertension was 31.5% among the weavers and 53.3% were prehypertensive. Both SBP (R 0.320, p-value <0.01) and DBP (R 0.366, p-value <0.01) had a significant positive correlation with the duration of exposure. Linear regression also significantly predicted SBP (p-value <0.01) and DBP (p-value <0.01) for the duration of noise exposure. In multinomial regression, for each year of exposure, the odds of hypertension increase by 10% (AOR 1.1, p-value <0.05) after adjusting for BMI, age, and smoking. Herein, we were able to demonstrate that duration noise exposure is independently associated to elevated blood pressure and increases the odds of hypertension.

## 1 Introduction

Hypertension, characterized by consistently high blood pressure, is a formidable global health challenge with far-reaching implications. The surge in cases over the past few decades, affecting 1.3 billion individuals worldwide, underscores the urgency of addressing this prevalent and life-threatening condition (1). Its impact extends beyond traditional health concerns, linking to various diseases affecting the heart, brain, kidneys, and even certain cancers (2–4). As a major contributor to non-communicable diseases (NCDs), hypertension represents 13% of total global deaths, emphasizing its role as a significant public health issue (5).

In the context of Bangladesh, the prevalence of hypertension is reported at 23.5% according to STEPS 2022, with variations in findings suggesting the need for comprehensive studies (6). Surprisingly, undiagnosed hypertension is as prevalent as diagnosed cases, and pre-hypertension, affecting a considerable portion of young adults, poses health risks comparable to hypertension itself (7,8). The economic burden of hypertension in Bangladesh is noteworthy, underscoring the multifaceted impact of this condition on both health and society (7,9).

Understanding the risk factors associated with hypertension is critical for effective prevention and management. Factors such as advancing age, family history, obesity, high-salt diets, physical inactivity, and excessive alcohol intake have been identified as contributors of hypertension (10,11). Additionally, environmental factors, including noise exposure, have been linked to hypertension and cardiovascular changes (12,13).

Occupational noise, prevalent in Bangladesh, particularly in industries like power looms, has been associated with cardiovascular problems. Power looms, as high-intensity noise-generating technologies, play a crucial role in Bangladesh’s economy, particularly in employing in rural areas (14). The association between occupational noise and arterial hypertension is well-established, with different types of occupations presenting varying degrees of risk (15).

Despite this, the specific relationship between the duration of occupational noise exposure and blood pressure remains understudied in Bangladesh. This research aims to bridge this gap by exploring the potential influence of the duration of noise exposure on the blood pressure of rural adult weavers in Bangladesh. With the weaving industry integral to the local economy and a significant proportion of the population engaged in it, understanding the nuanced dynamics between occupational noise exposure and cardiovascular health becomes imperative.

The study, conducted in the greater area of Narayanganj, focuses on weaving factories that produce fabric from threads in power looms. These machines generate continuous and extreme mechanical sound, potentially exceeding the threshold limit value (TLV) of noise exposure set by NIOSH (The National Institute for Occupational Safety and Health). Previous research in different countries has reported high noise levels in similar industries, emphasizing the need for a comprehensive examination in the Bangladeshi context (15, 16).

This research will contribute valuable insights into the impact of prolonged noise exposure on blood pressure levels, addressing a discernible gap in the existing literature. By focusing on rural adult weavers and collecting raw data from the research area, the study aspires to enhance our understanding of the intricate dynamics between occupational noise exposure and cardiovascular health. The findings are expected to provide a nuanced perspective on the relationship between the duration of noise exposure and blood pressure, thus making a valuable contribution to the scientific discourse on this subject.

## 2 Article types

Original research article

## 3. Materials and Methods

### 3.1 Study population and data source

This was a cross-sectional study conducted among rural adult loom weavers working in power-loom factories in Narayanganj, Bangladesh from 1st January 2023 to 31st December 2023. Data were collected from selected power-loom factories in Gazipura village, Araihazar. The sample size was determined by assuming the prevalence of hypertension in adult population of Bangladesh (STEPS 2022), 95% confidence limit, and 5% marginal error, and the sample size was 276. Participants were selected by convenience sampling technique and data were collected following through face-to-face interviews using a semi-structured questionnaire and physical measurement. The questionnaire was pre-tested on 10% of the total sample in Bishnondi, Araihazar, Narayongonj, Bangladesh. Weavers, actively working in the factories, were included in the study. Those who had career breaks or interruptions and females who were pregnant have been excluded from this study.

### 3.2 Assessment of variables

Tools were a digital weight machine for weight measurement, a non-stretchable tap for height measurement, a sphygmomanometer and stethoscope for blood pressure management, and NIOSH SLM app in iPhone 11 for noise measurement. Data was collected by the researcher using KOBO toolbox in a Samsung s6 lite tab. For assessing nutritional status, the Asian classification of BMI was used (33)

Blood pressure was measured using an ALPK2 ANEROID sphygmomanometer with an adult cuff-size and M stethoscope. Participants were instructed to sit quietly in a selected insulated place and rest for 10 minutes before the measurement. The measurement was taken on the left arm with the participant’s forearm resting on a table, palm facing upward. Two readings were taken at a 3-minute interval. If the difference between the first two readings exceeded 5 mmHg, a third reading was recorded. The mean of the two or three readings was calculated. Systolic and diastolic blood pressure readings were rounded up to the nearest 5 units and recorded. Data collection and physical measurements took place while the workers were in their off-duty period.

Sound intensity within the factories was assessed using the NIOSH SLM app installed on an iPhone 11. R^2^ of measurements in NIOSH SLM while using an iPhone and calibrated pure tone sound field was 0.97 (32). The measurements were conducted for 15 minutes at various points inside the factories, specifically near the machines where the workers were stationed.

### 3.3 Ethical considerations

Ethical approval was granted by the ethics committee of National Institute of Preventive and Social Medicine (approval number NIPSOM/IRB/2023/06).

### 3.4 Data analysis

After proper verification, the data Excel sheet was entered into SPSS window version 23.0. After entries, data consistency was checked. Data were analyzed according to the objectives of the study. Descriptive statistics were used to describe the variables in percentage, mean, and SD, as appropriate. The analysis also included correlation to test the association between various variables. *t-test* was done between different variables. The χ2 test was also used to test the association between attributes. Linear regression was used to predict the association between systolic blood pressure, diastolic blood pressure, and duration of noise exposure. Multinomial logistic regression was done to look for association between duration of exposure and blood pressure categories controlling other confounders.

## 4 Results

### 4.1 Socio-demographic characteristics of workers

In this cross-sectional study, the research focused on 289 rural adults employed as power-loom weavers. The average age of the respondents was 34.41(±10.17) years. A significant majority, comprising over three-fourths of the participants, fell below the age of 40, whereas only 7.3% were aged 50 or above. Among the respondents, the male population dominated, constituting the majority at 94.1%, while only 5.9% were females.

A substantial percentage 84.4% were married. Unmarried individuals accounted for 12.1% of the total, while the remaining respondents were either divorced or widowed. Almost the entire respondent group, except a single individual, identified as Muslims.

The highest proportion, comprising 42.9%, had completed primary education. In comparison, 36.7% reported being without formal education, and 19.0% had attained a secondary education. A minimal 1.4% had achieved a higher level of education.

In terms of income, the mean family income was 36,747.42 taka (±12,300), while the mean personal income was 18,257.44 taka (±4,281.79). Approximately 31.5% of the respondents were aware of family members having or having had hypertension, while 15.6% were certain of its absence. Furthermore, 18% and 20.1% reported the presence of cardiovascular disease and diabetes, respectively, within their families. Only a small fraction of respondents was knowledgeable about their comorbidities, such as cardiovascular diseases (6.6%) or diabetes mellitus (4.5%).

### 4.2 Occupational attributes related to noise

Workers have been working for a mean duration of 15.97 years in the power loom factories. The majority were found to be working for 11-20 years in this weaving industry. The noise level, measured during the working period in the factories was on average 105.45 dB, with a minimum intensity of 96 dB and a Maximum of 111 dB.

### 4.3 Health behavior

This study found that 52% of participants had regular smoking habits, while 38.5% were nonsmokers. Only 7% acknowledged regular consumption of smokeless tobacco, while the majority abstained from it. The mean daily servings of fruits and vegetables were 1.89, with 84.4% limiting fruit intake to a single serving. About 42% had a history of alcoholic drinks. Table salt usage varied, with 58.8% reporting regular use and 17.6% never using it. 81% reported frequent soft drink consumption, 61.6% consuming energy drinks, and 57.1% consuming both. 50.5% of the population did not engage in moderate to vigorous physical activity. The average BMI was 21.69, with underweight being most prevalent in the younger age group and obesity increasing with age. Normal BMI had an inverse relationship with overweight across all age groups.

#### 4.4 Blood pressure and association with factors

Among the respondents, only 15.2% were found to have normal blood pressure, whereas 31.5% (91 respondents) were diagnosed with hypertension, including those with a prior diagnosis. Most respondents (53.3%) fell into the prehypertension category. Within the hypertensive group (91 respondents), only 35.2% were aware of their condition, while 64.8% (59 respondents) were unaware of their elevated blood pressure.

For SBP, the mean was 124.07 mmHg for males and 131.76 mmHg for females. Regarding DBP, the mean was 82.72 mmHg for males and 86.44 mmHg for females. The overall mean blood pressure, combining both male and female data, was 124.52 mmHg for SBP and 82.93 mmHg for DBP.

The duration of work showcased a positive correlation coefficient of 0.320 (p <0.01) with systolic blood pressure. The duration of work revealed also a notable positive correlation of 0.366 (p <0.01), the positive correlations suggest that as the duration of work increases, there is a corresponding increase in both systolic and diastolic blood pressure levels.

A linear regression analysis was conducted to examine the relationship between systolic blood pressure and the duration of exposure in male respondents, controlling for factors like BMI, age, physical activity, and smoking. The results showed a significant increase in systolic blood pressure by 0.574 mmHg for each year of exposure, and 0.411 mmHg in diastolic blood pressure. A multinomial logistic regression was performed to determine the association between blood pressure categories (normal, prehypertension, hypertension) and the duration of noise exposure. The model included four independent variables, including duration of noise exposure, age, BMI, and smoking status. The adjusted odds ratio of workplace noise exposure with hypertension was found to be 1.1, indicating a 10% increase in hypertension with each year of exposure. However, the association between prehypertension and noise exposure was insignificant.

## 5 Discussion

In this study, a sample of 289 individuals was analyzed, primarily young adults (20-40 years) with a mean age of 43.41 years. The majority were males (94.1%), reflecting the gender distribution in power loom factories. The mean duration of workplace noise exposure was 15.97 years, where the noise level exceeded the recommended threshold limit value (TLV) at a mean noise level of 105.45 dB (16–18).

Regarding blood pressure, it was observed that, mean SBP of 124.52 mmHg and DBP of 82.93 mmHg among those exposed to noise exceeding 85dB. Compared to general population surveys, this study indicated higher blood pressure levels, with 31.5% of participants classified as hypertensive, while in Bangladesh, according to the STEPS survey, it was 23.5% (20). This prevalence surpassed findings in India and China, suggesting a potentially unique impact of noise exposure in our studied population (17,19).

The study explored correlations between various factors and blood pressure levels. Age demonstrated a moderate positive correlation with both SBP (0.345) and DBP (0.335), aligning with previous research findings (21, 22). Additionally, we noted differences in blood pressure between genders, with females exhibiting higher mean SBP (131.76 mmHg) and DBP (86.44 mmHg) compared to males. This contrasts with studies in China and supports the notion that occupational roles may influence these gender differences (19).

Smoking prevalence among participants was 52%, contradicting national averages, and presented complex relationships with blood pressure. While smoking in this study did not follow the expected pattern of increased blood pressure, the intricate interplay of smoking, BMI, and blood pressure aligns with existing literature (23–25).

Physical activity emerged as a significant factor, with inactive individuals having higher mean SBP (123.02 mmHg) and DBP (81.52 mmHg) compared to those engaging in physical activities. The study revealed elevated noise levels in textile factories (105.45 dB), potentially contributing to increased blood pressure. Notably, none of the participants used safety earplugs, a practice that could significantly reduce noise exposure (26).

The investigation revealed a noteworthy correlation (r = 0.320, p < 0.01) between the duration of occupational noise exposure and systolic blood pressure, suggesting that an increase in exposure duration corresponds to elevated systolic blood pressure. A similar pattern was observed for diastolic blood pressure, with a slightly higher correlation coefficient of 0.366 (p < 0.01). These findings underscore a direct relationship between prolonged occupational noise exposure and the development of hypertension in weavers.

In a study of Indonesian textile weavers, exposure exceeding 10 years was associated with significantly higher systolic and diastolic blood pressure, with odds ratios of 3.538 (p < 0.05) and 3.123 (p < 0.05), respectively (26). Another investigation noted elevated mean systolic and diastolic blood pressure in workers with more than two years of exposure compared to those with less than two years (27).

Multinomial regression analysis, adjusting for BMI, age, and smoking status, further substantiated the association between the duration of noise exposure and an increased risk of hypertension among workers. Specifically, each year of exposure was associated with 10% higher odds of developing hypertension in the study area. While prehypertension had an adjusted odds ratio (AOR) of 1.058, it did not reach statistical significance. A recent study indicated a similar trend, reporting increased prehypertension rates in a noisy industry, with AORs of 2.8 for 6-10 years’ experience and 4.8 for more than 10 years (27).

Consistent with these findings, studies among Indian weavers demonstrated a significant prevalence of hypertension among those with over 10 years of experience (128). Studies conducted by Wang et al. and Sbihi, Davies, and Demers also linked increased rates of hypertension to the growing duration of occupational noise exposure (19,29). Moreover, a study in Brazil highlighted that exposure to occupational noise levels exceeding 85 dB(A) for over 10 years was associated with high blood pressure among oil industry workers (30). A systematic review encompassing 50 studies emphasized a considerable prevalence of hypertension among cotton workers in low-and middle-income countries, considering the duration of noise exposure (31). These collective findings contribute significantly to the mounting evidence supporting the substantial association between prolonged noise exposure and an increased risk of hypertension among workers.

## 6 Conclusion

This study concludes that prolonged exposure to high-intensity noise in textile mills is associated with elevated blood pressure among workers, increasing the risk of cardiovascular diseases, particularly hypertension. To mitigate these risks, routine blood pressure monitoring is recommended, alongside the provision of noise-canceling ear protection for weavers. Additionally, monitoring and addressing lifestyle factors like smoking and alcohol consumption are crucial. The study emphasizes the need for ongoing research with larger sample sizes to further confirm and expand upon these findings, contributing to the development of effective health strategies for textile workers.

**Figure 1.**
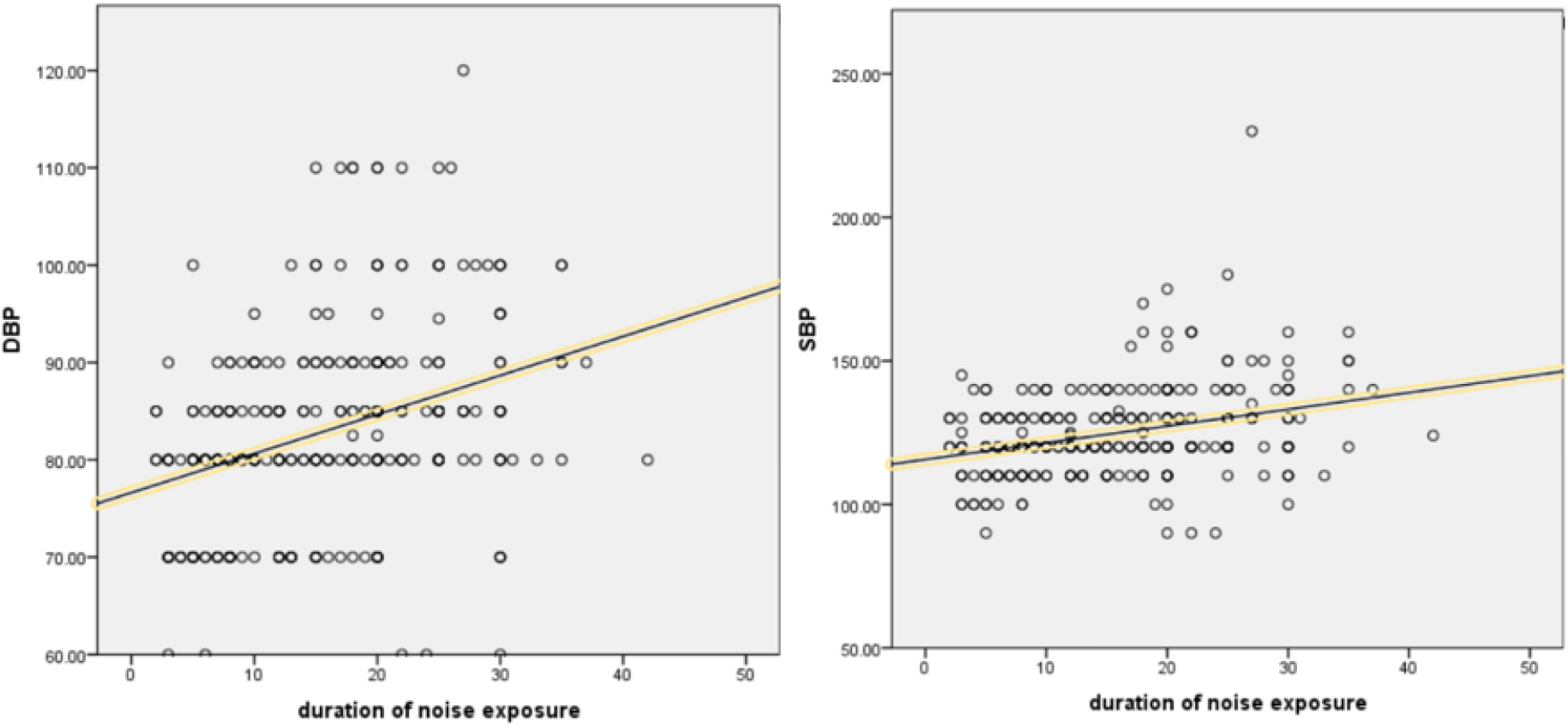
Correlation of SBP and DBP with duration of noise exposure

**Table 1.** Sociodemographic characteristics of the respondents.

| **Characteristics** | **Frequency (n)** | **Percentage (%)** |
| --- | --- | --- |
| **Age (in years)** |  |  |
| 11-20 | 21 | 7.3 |
| 21-30 | 94 | 32.5 |
| 31-40 | 107 | 37 |
| 41-50 | 46 | 15.9 |
| More than 50 | 21 | 7.3 |
| **Sex** |  |  |
| Male | 272 | 94.1 |
| Female | 17 | 5.9 |
| **Education Level** |  |  |
| No formal education | 106 | 36.7 |
| Primary | 124 | 42.9 |
| Secondary | 55 | 19.0 |
| Higher secondary or above | 4 | 1.4 |
| **Type of Family** |  |  |
| Joint family | 81 | 28 |
| Nuclear family | 208 | 72 |
| **Type of Residence** |  |  |
| Brick built | 9 | 3.1 |
| Tin shade | 59 | 20.4 |
| Tin built house | 221 | 76.5 |
| **Monthly Family Income (in BDT)** |  |  |
| <20000 | 147 | 50.9 |
| 20000-50000 | 130 | 45 |
| More than 50000 | 12 | 4.2 |

**Table 2.** Health behavior of respondents.

|  | Frequency | Percentage (%) |
| --- | --- | --- |
| **Smoking tobacco products** |  |  |
| Regular smoker | 151 | 52.4 |
| Occasional smoker | 15 | 5.2 |
| Past smoker | 11 | 3.8 |
| Non-smoker | 111 | 38.5 |
| **Smokeless tobacco product** |  |  |
| Regular user | 20 | 7 |
| Occasional user | 92 | 32.2 |
| Past user | 13 | 4.5 |
| Non-user | 161 | 56.3 |
| **Alcohol consumption** |  |  |
| Ever in life | 122 | 42.2 |
| Never | 167 | 57.8 |
| **Use of table salt** |  |  |
| Always | 170 | 58.8 |
| Often | 24 | 8.3 |
| Sometimes | 24 | 8.3 |
| Rarely | 20 | 6.9 |
| Never | 51 | 17.6 |
| **Carbonated drinks** |  |  |
| Soft drinks | 234 | 81 |
| Energy drinks | 178 | 61.6 |
| Both | 165 | 57.1 |

**Table 3.** Multinomial regression.

| Characteristics | Blood pressure category** | coefficient | AOR | 95% CI for AOR |  | p-value |
| --- | --- | --- | --- | --- | --- | --- |
|  |  |  |  | Lower | Upper |  |
| Age | Prehypertension | .007 | 1.007 | .951 | 1.066 | .949 |
|  | Hypertension | .045 | 1.046 | .984 | 1.112 | .146 |
| BMI | Prehypertension | .046 | 1.047 | .931 | 1.177 | .442 |
|  | Hypertension | .214 | 1.239 | 1.085 | 1.414 | <0.05 |
| Duration of exposure | Prehypertension | .057 | 1.058 | .988 | 1.133 | .105 |
|  | Hypertension | .095 | 1.100 | 1.022 | 1.184 | <0.05 |
| Smoking status |  |  |  |  |  |  |
| Regular smoker | Prehypertension | -.810 | .445 | .195 | 1.015 | .054 |
|  | Hypertension | -1.567 | .209 | .083 | .523 | .001 |

## Data Availability

All data produced in the present study are available upon reasonable request to the authors

## 10 Author Contributions

GDP and MRP designed the study. GDP collected the data. GDP, MRP, SA performed statistical analysis. GDP, RT and OIR drafted this manuscript. SA, DJ, SUA and RT revised the manuscript. GDP had primary responsibility for final content. All authors were involved in the revisions and approved the final version of the manuscript.

## 12 Acknowledgments

We thank the power-loom weavers of Araihajar for their contributions to this project, and express our sincere support and gratitude to them.

## 14 Data Availability Statement

The raw data supporting the conclusions of this article will be made available by the authors, without undue reservation.

